# The prevalence of olfactory dysfunction and its associated factors in patients with COVID-19 infection

**DOI:** 10.1101/2021.01.27.21250153

**Authors:** Md Mehedi Hasan, Naima Ahmed Tamanna, Mohammad Nasimul Jamal, Md Jamal Uddin

**Affiliations:** Specialist, Department of ENT & Head Neck Surgery, Square Hospitals Ltd., Dhaka, Bangladesh; Research student, Department of Statistics, Jagannath University, Dhaka, Bangladesh; Consultant, Department of ENT & Head Neck Surgery, Square Hospitals Ltd., Dhaka, Bangladesh; Department of Statistics, Shahjalal University of Science and Technology, Sylhet, Bangladesh

**Author notes:** Corresponding author’ s address, Md Mehedi Hasan, Department of ENT & Head Neck Surgery, 18/F, Bir Uttam Qazi Nuruzzaman Sarak, West Panthapath, Dhaka-1205. Funding: None. Conflicts of interest: The authors have no conflicts of interest. Authors contribution: Md Mehedi Hasan: Design, collect and entry data, draft the manuscript and review the manuscript. Naima Ahmed Tamanna: Organize the results, Draft the manuscript and review the manuscript. Mohammad Nasimul Jamal: Provide editorial comments and review the manuscript. Md Jamal Uddin: Analysis the data and review the manuscript.

**Keywords:** COVID-19, Anosmia, Olfactory Dysfunction, Factors, Bangladesh

## Abstract

**Objective:** To determine the prevalence of olfactory dysfunctions, mainly, anosmia and to identify its associated factors in patients with COVID-19 infection.

**Study design:** A hospital-based prospective observational cohort study

**Setting:** A COVID dedicated hospital, Square Hospitals Ltd., Dhaka, Bangladesh.

**Methods:** We collected patients’ information including laboratory-confirmed COVID-19 test results. We used Pearson Chi-square test and logistic regression model to assess the associations between demographic and clinical characteristics and olfactory outcomes.

**Results:** Out of 600 COVID-19 positive patients, 38.7% were diagnosed with olfactory dysfunction. Our analyses showed that patients’ age, smoking status, cough, dyspnea, sore throat, asthenia, and nausea or vomiting were significantly associated with the anosmia. We observed the risk of developing anosmia was greater in younger patients than in older patients, and this risk decreased as age increased [odds ratio (OR) range for different age groups: 1.26 to 1.08]. Smoking patients were 1.73 times more likely to experience anosmia than non-smoking patients [OR=1.73, 95% confidence interval (CI) = 1.01-2.98]. In addition, patients complained asthenia had a significantly double risk of developing the anosmia [OR = 1.96, CI = 1.23-3.06].

**Conclusions:** Our study shows that about 39% of patients diagnosed with olfactory dysfunction. Patients’ age, smoking status, and asthenia are significantly positively associated with the anosmia. Since anosmia can be a significant marker for the diagnosis of COVID-19, we suggest regular screening of olfactory dysfunction in patients with early symptoms of COVID-19, particularly younger patients, smoker, and complained asthenia.

## Introduction

The 2019 coronavirus disease (COVID-19) is an ongoing viral pandemic that has arisen from East Asia (China) and is rapidly spreading to every corner of the world.^1^ COVID-19 was confirmed a global pandemic by the World Health Organization (WHO) on March 11, 2020. As of January 20, 2021, there are 96,281,077 identified cases and 2,056,521 deaths of COVID-19 worldwide.^2^

The virus is transmitted from human to human by the transmission of droplets and direct contact with the mucous membranes of oral, nasal, and eye.^3^ The most common symptoms of the COVID-19 patients are fever, dry cough, sore throat, fatigue, dyspnea and so on.^4^ Moreover, olfactory or gustatory dysfunction (e.g. anosmia-temporary loss of smell) is often considered a symptom of COVID-19.^4 5 6 7^ Several studies have also documented a high incidence of olfactory dysfunction, especially anosmia, in patients with COVID-19, often considered to be the first symptom.^5 8 9 10 11 12 13 14^ Anosmia is defined as the range of olfactory dysfunction (OD), which can be caused by a variety of causes such as, allergic, nasal polyps, viral illnesses, upper respiratory tract infections etc. ^15 16^ Furthermore, Brann et al. clarified that an inflammation of non-neuronal support cells in the nose and forebrain of COVID-19 patients could be responsible for an anosmia.^17^ They also noted that, nasal obstruction is not related with anosmia in this case; thus, such patients might experience anosmia without complaining stuffy nose. However, it is unclear which types of COVID-19 patients are mostly getting this symptom.

As far we know, there is no study placed about olfactory dysfunction for COVID-19 infected patients and its associated factors in Bangladesh. We aimed to determine the correlation between OD, particularly anosmia and COVID-19 infected patients’ demographic and clinical characteristics.

## Methods

### Ethical statement and inform consent

Informed consent was taken from the patient or legal guardian of the patient. Moreover, the aims and objectives of the study, along with its procedure, risk and benefit were explained to the patients.

### Ethics committee approval

The ethical clearance was taken from the ethical committee at SQUARE Hospital Ltd.

### Subjects and setting

We conducted a hospital-based prospective observational study from July 2020 to September 2020. We collected clinical data of patients with laboratory-confirmed COVID-19 infection from a tertiary level COVID dedicated hospital, name: Square Hospitals Ltd., Dhaka, Bangladesh.

We considered several inclusion and exclusion criteria. The inclusion criteria were: laboratory-confirmed COVID-19 infection (reverse transcription-polymerase chain reaction, RT-PCR); adult (> 18 years old); both male and female; and patients clinically stable to answer the questionnaire. Moreover, the exclusion criteria were: patients with olfactory dysfunctions before February 2020; patients without a finding of laboratory-confirmed COVID-19 infection; patients in the intensive-care unit (ICU) at the time of the study (due to the restrictions from the hospital & their clinical condition). Accordingly, we mostly included mild-to-moderate COVID-19 patients without the need for ICU.

### Clinical outcomes

Clinical data have been recorded during the ear, nose, and throat (ENT) consultation prospectively; in the patients’ room who were admitted in the hospital; or over the phone for infected health professionals. The questionnaire consisted of some general questions, such as age, sex, ethnicity, education, occupation, smoking or other drug addiction; three general clinical questions (e.g., comorbidities such as diabetes, hypertension, chronic kidney disease (CKD), Ischemic heart disease (IHD), asthma, thyroid, dyslipidemia, and cancer; general symptoms and ENT symptoms related with COVID-19 infection); and one question about the treatment of the COVID-19 infection. All patients were requested to complete the questionnaire in the presence of the investigator.

### Olfactory outcomes

The questionnaire has identified the incidence of anosmia. The questions have been selected to illustrate the timing and accompanying symptoms of olfactory dysfunctions. The patients who lost their sense of smell during COVID-19 were identified as anosmic patients. These anosmic patients had no previous history of olfactory dysfunction or any surgery or trauma within two months before COVID-19. It is to clarify that we evaluated the mean recovery time of olfaction with four defined hypotheses: 1–4 days; 5–8 days; 9–14 days; and >15 days. Mentioning to the studies demonstrate that the viral load was significantly decreased after 14 days and could recover within 28 days.^16^

### Statistical methods

The probable associate factors between epidemiological, clinical, and olfactory outcomes have been measured by using the Pearson Chi-square test. Partial or unfinished responses were excluded from the analysis. A level of p < 0.05 was used to determine statistical significance. As the outcome variable anosmia was a binary variable, we applied a binary logistic regression model to identify the association between patients’ Anosmia and essential characteristics. Statistical Package for the Social Sciences for Windows (SPSS version 25) was used to perform the statistical analyses.

## Results

A total of 600 COVID-19 infected patients participated in the study, and 232 (38.7%) patients were diagnosed with olfactory dysfunction. Of the olfactory dysfunction patients, 34% had this symptom for 5 to 8 days, followed by 30% for 11 to 14 days (Figure 1). There were 383 (64%) males and 217 (36%) females. Also, among the patients with anosmia, 62.1% were male (Figure 2). The mean age of patients was 50.20 ± 14.81 years (range: 22-88 years). The highest percentage (33.6%) of anosmia patients belong to the age group 30-39 years. 75.4% were from the urban area, 62.9% completed a university degree. Among all patients, 62% had comorbidities, and out anosmia patients, 51.3% had comorbidity where the most prevalent comorbidities of patients were diabetes and hypertension (Figure 3). As expected, the highest percentage (75%) of patients had a fever, and the lowest parentage (3.9%) of patients had nausea or vomiting (Figure 4). From bivariate analyses, we observed that age (P<0.001), education (P<0.001), diabetes (P=0.001), hypertension (P<0.001), and CKD (ESRD) (P=0.025) were significantly associated with the anosmia. We also showed that most of the symptoms were also significantly related to the Anosmia (Table-1).

**Figure 1.**
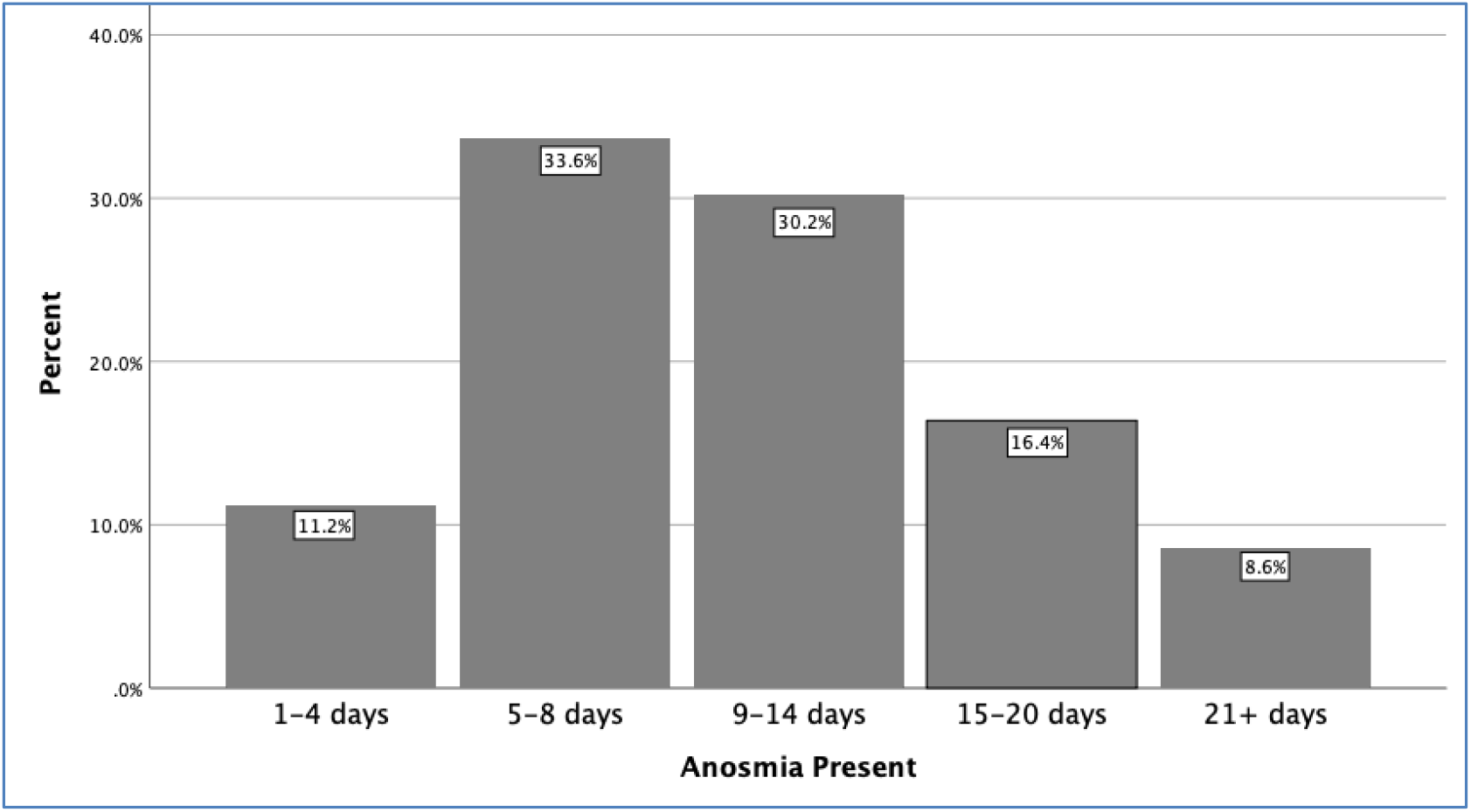
Percentage of COVID-19 patients with duration anosmia present

**Figure 2.**
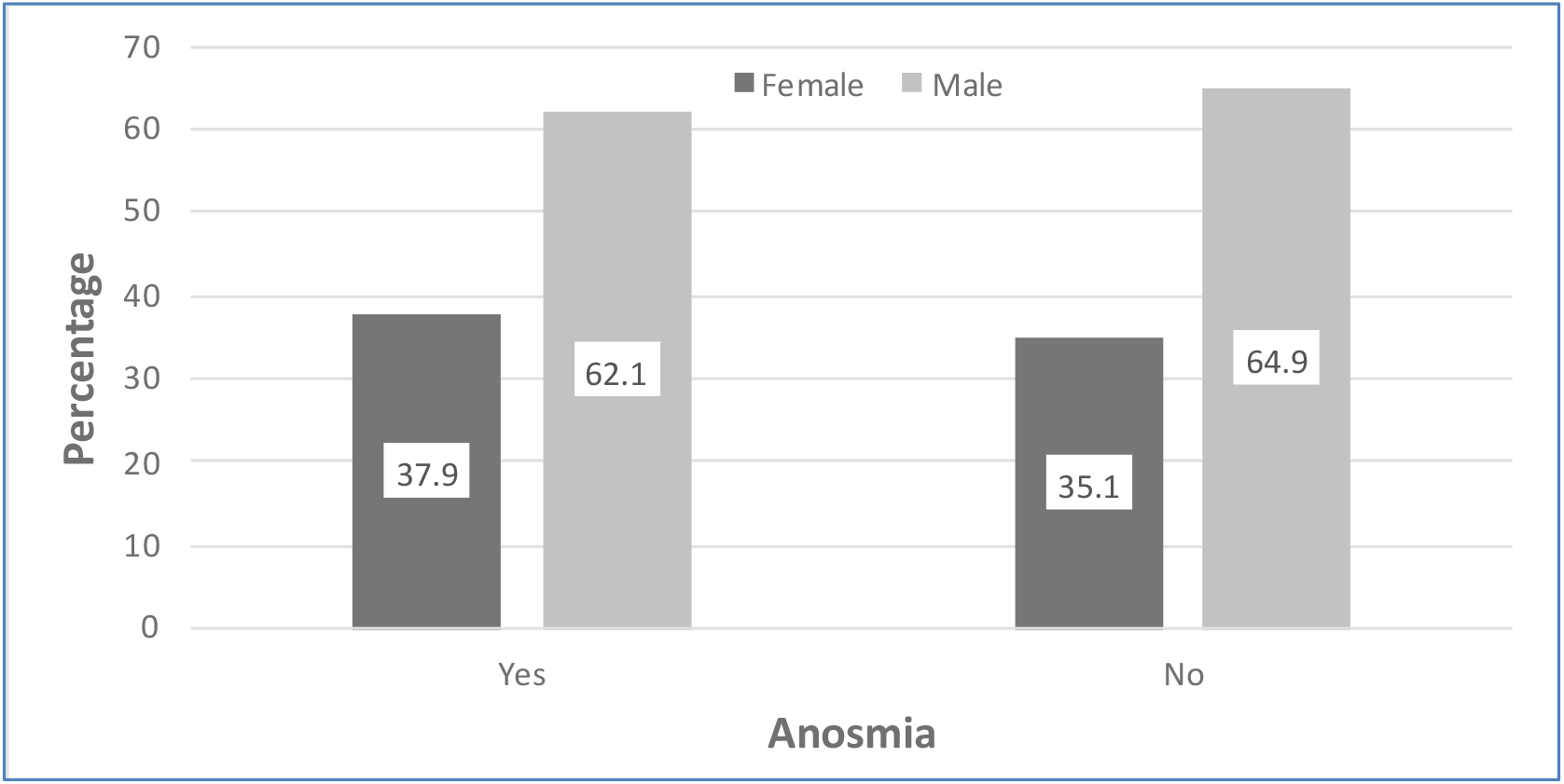
Percentage of male and female in COVID-19 patients with Anosmia

**Figure 3.**
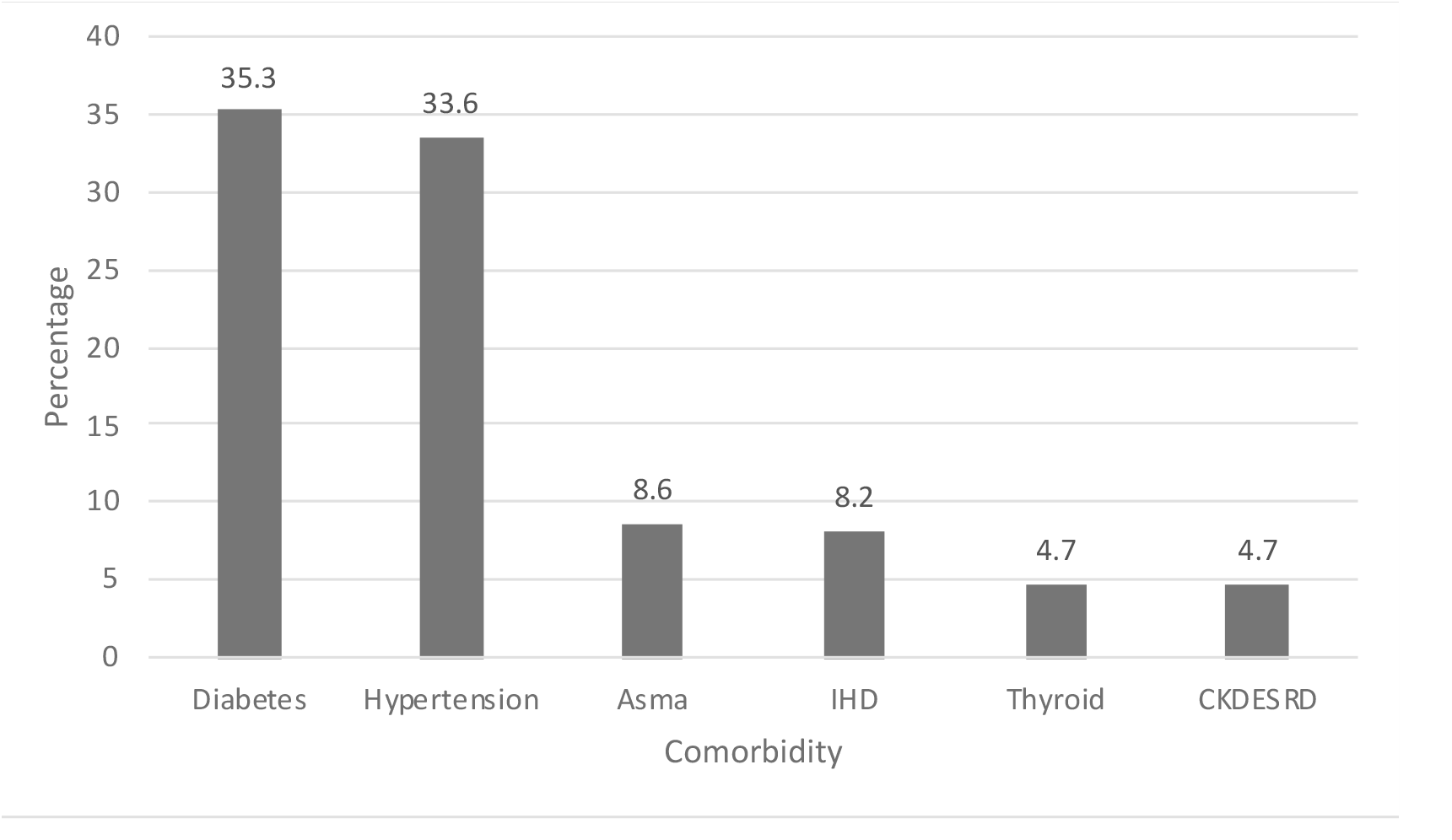
Percentage of different comorbidities in COVID-19 patients with Anosmia

**Figure 4.**
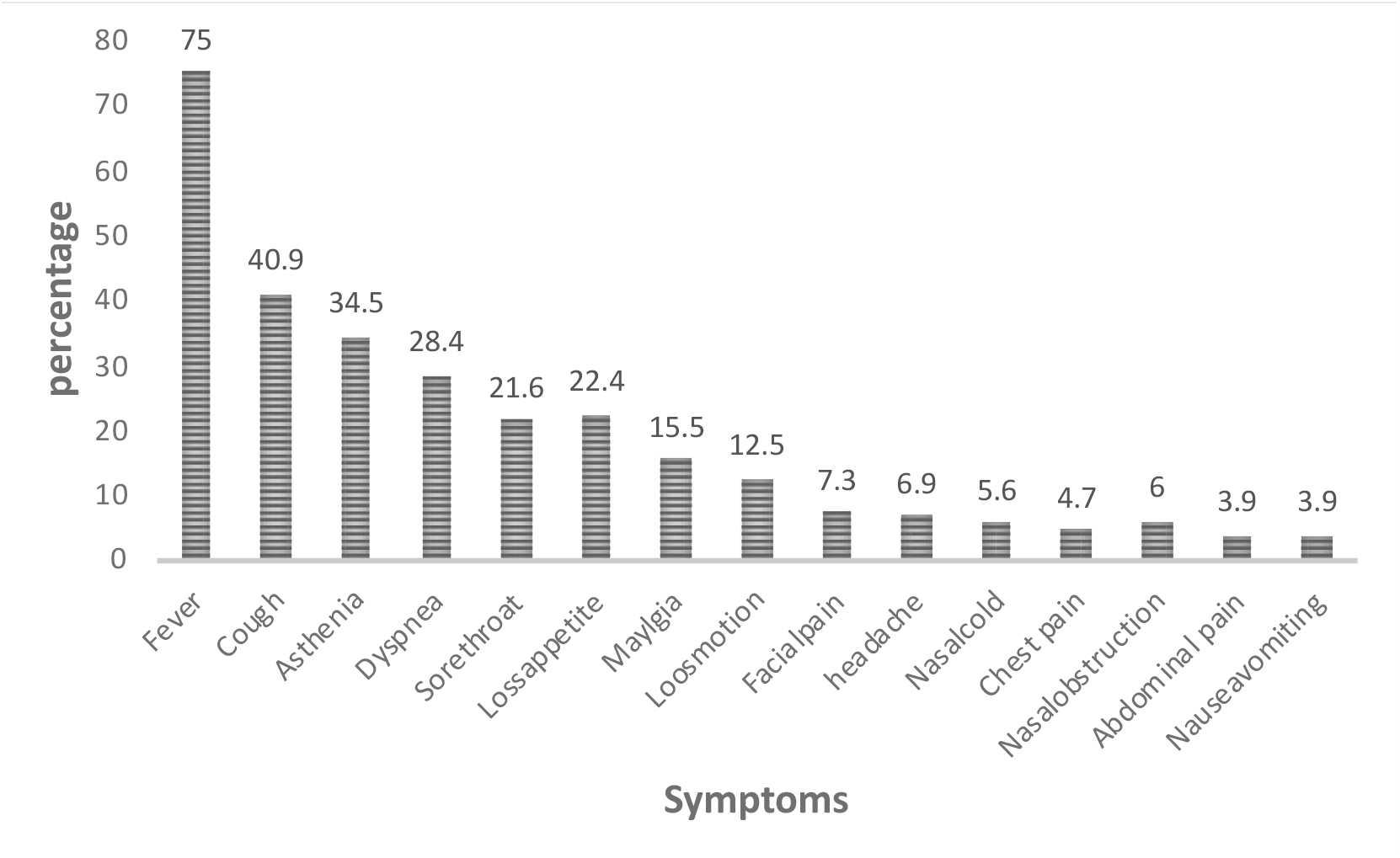
Percentage of different symptoms in COVID-19 patients with Anosmia

**Table 1.** Distribution of patients’ socio-demographic and clinical characteristics across the outcome variable (anosmia) including p-value from Pearson Chi-Square Tests.

|  |  | Anosmia |  |  |  | P-value |
| --- | --- | --- | --- | --- | --- | --- |
|  |  | No |  | Yes |  |  |
|  |  | Count | Column % | Count | Column % |  |
| Gender | Female | 129 | 35.1 | 88 | 37.9 | 0.475 |
|  | Male | 239 | 64.9 | 144 | 62.1 |  |
| Age (in years) | 20-29 | 16 | 4.3 | 17 | 7.3 | 0.000 |
|  | 30-39 | 69 | 18.8 | 78 | 33.6 |  |
|  | 40-49 | 62 | 16.8 | 46 | 19.8 |  |
|  | 50-59 | 90 | 24.5 | 32 | 13.8 |  |
|  | 60+ | 131 | 35.6 | 59 | 25.4 |  |
| Area | Rural | 81 | 22.0 | 57 | 24.6 | 0.468 |
|  | Urban | 287 | 78.0 | 175 | 75.4 |  |
| Education | Post-graduation | 34 | 9.2 | 34 | 14.7 | 0.000 |
|  | Graduation | 186 | 50.5 | 146 | 62.9 |  |
|  | Higher Secondary | 83 | 22.6 | 39 | 16.8 |  |
|  | Secondary School | 44 | 12.0 | 8 | 3.4 |  |
|  | Primary | 21 | 5.7 | 5 | 2.2 |  |
| Diabetic | No | 187 | 50.8 | 150 | 64.7 | 0.000 |
|  | Yes | 181 | 49.2 | 82 | 35.3 |  |
| Hypertension | No | 177 | 48.1 | 154 | 66.4 | 0.000 |
|  | Yes | 191 | 51.9 | 78 | 33.6 |  |
| Asthma | No | 324 | 88.0 | 212 | 91.4 | 0.197 |
|  | Yes | 44 | 12.0 | 20 | 8.6 |  |
| Thyroid | No | 350 | 95.1 | 221 | 95.3 | 0.934 |
|  | Yes | 18 | 4.9 | 11 | 4.7 |  |
| Dyslipidemic | No | 344 | 93.5 | 225 | 97.0 | 0.059 |
|  | Yes | 24 | 6.5 | 7 | 3.0 |  |
| Depression | No | 365 | 99.2 | 231 | 99.6 | 0.059 |
|  | Yes | 3 | 0.8 | 1 | 0.4 |  |
| Chronic Kidney | No | 332 | 90.2 | 221 | 95.3 | 0.025 |
|  | Yes | 36 | 9.8 | 11 | 4.7 |  |

| **Disease** |  |  |  |  |  |  |
| --- | --- | --- | --- | --- | --- | --- |
| **IHD** | No | 322 | 87.5 | 213 | 91.8 | 0.098 |
|  | Yes | 46 | 12.5 | 19 | 8.2 |  |
| **Neurological Disease** | No | 357 | 97.0 | 231 | 99.6 | 0.029 |
|  | Yes | 11 | 3.0 | 1 | 0.4 |  |
| **Allergic Rhinitis** | No | 366 | 99.5 | 230 | 99.1 | 0.640 |
|  | Yes | 2 | 0.5 | 2 | 0.9 |  |
| **Cancer** | No | 360 | 97.8 | 230 | 99.1 | 0.222 |
|  | Yes | 8 | 2.2 | 2 | 0.9 |  |
| **Comorbidity** | No | 115 | 31.3 | 113 | 48.7 | 0.000 |
|  | Yes | 253 | 68.8 | 119 | 51.3 |  |
| **Smoking or Other Addiction** | No | 308 | 83.7 | 186 | 80.2 | 0.270 |
|  | Yes | 60 | 16.3 | 46 | 19.8 |  |
| **Fever** | No | 56 | 15.2 | 58 | 25.0 | 0.003 |
|  | Yes | 312 | 84.8 | 174 | 75.0 |  |
| **Cough** | No | 155 | 42.1 | 137 | 59.1 | 0.000 |
|  | Yes | 213 | 57.9 | 95 | 40.9 |  |
| **Dyspnea** | No | 216 | 58.7 | 166 | 71.6 | 0.001 |
|  | Yes | 152 | 41.3 | 66 | 28.4 |  |
| **Sore Throat** | No | 241 | 65.5 | 182 | 78.4 | 0.001 |
|  | Yes | 127 | 34.5 | 50 | 21.6 |  |
| **Asthenia** | No | 299 | 81.3 | 152 | 65.5 | 0.000 |
|  | Yes | 69 | 18.8 | 80 | 34.5 |  |
| **Myalgia** | No | 307 | 83.4 | 196 | 84.5 | 0.732 |
|  | Yes | 61 | 16.6 | 36 | 15.5 |  |
| **Loss Appetite** | No | 319 | 86.7 | 180 | 77.6 | 0.004 |
|  | Yes | 49 | 13.3 | 52 | 22.4 |  |
| **Loose Motion** | No | 315 | 85.6 | 203 | 87.5 | 0.509 |
|  | Yes | 53 | 14.4 | 29 | 12.5 |  |
| **Headache** | No | 326 | 88.6 | 216 | 93.1 | 0.068 |
|  | Yes | 42 | 11.4 | 16 | 6.9 |  |
| **Nausea or Vomiting** | No | 334 | 90.8 | 223 | 96.1 | 0.013 |
|  | Yes | 34 | 9.2 | 9 | 3.9 |  |
| **Nasal Cold** | No | 336 | 91.3 | 219 | 94.4 | 0.161 |
|  | Yes | 32 | 8.7 | 13 | 5.6 |  |
| **Facial Pain** | No | 346 | 94.0 | 215 | 92.7 | 0.514 |
|  | Yes | 22 | 6.0 | 17 | 7.3 |  |
| **Abdominal Pain** | No | 346 | 94.0 | 223 | 96.1 | 0.258 |
|  | Yes | 22 | 6.0 | 9 | 3.9 |  |
| **Chest Pain** | No | 353 | 95.9 | 221 | 95.3 | 0.697 |
|  | Yes | 15 | 4.1 | 11 | 4.7 |  |
| **Nasal Obstruction** | No | 340 | 92.4 | 218 | 94.0 | 0.462 |
|  | Yes | 28 | 7.6 | 14 | 6.0 |  |

Table-2 shows the association between patients’ characteristics and anosmia from multivariable binary logistic regression model. We found that patients’ age, smoking status, cough, dyspnea, sore throat, asthenia, and nausea vomiting were significantly associated with the anosmia. The risk of developing anosmia for younger patients was higher than the older patients, and this risk decreased with the increasing age [OR 1.26 to 1.08]. Patients with smoking or other addiction had a 1.73 times higher chance to develop anosmia than non-smoker [OR=1.73, 95% confidence interval (CI) = 1.01-2.98]. Besides, patients complained asthenia had significantly double risk to develop the anosmia [OR = 1.96, CI = 1.23-3.06, p =0.001]. All other statistically significant variables showed negative relation with the anosmia.

**Table 2.** Association between patients’ characteristics and Anosmia from multivariable logistic regression model.

| Factors | Categories | Estimate | OR | 95% C.I. for OR |  | P-value |
| --- | --- | --- | --- | --- | --- | --- |
|  |  |  |  | Lower | Upper |  |
| **Gender** | Female | 0.16 | 1.17 | 0.77 | 1.80 | 0.46 |
|  | Male (Ref) |  |  |  |  |  |
| **Age (in years)** | 20-29 | 0.23 | 1.26 | 0.52 | 3.06 | 0.61 |
|  | 30-39 | 0.24 | 1.27 | 0.70 | 2.32 | 0.43 |
|  | 40-49 | 0.08 | 1.08 | 0.60 | 1.96 | 0.79 |
|  | 50-59 | -0.59 | 0.55 | 0.31 | 0.99 | 0.05 |
|  | 60+ (Ref) |  |  |  |  |  |
| **Diabetic** | Yes | 0.11 | 1.12 | 0.69 | 1.80 | 0.65 |
|  | No (Ref) |  |  |  |  |  |
| **Hypertension** | Yes | -0.40 | 0.67 | 0.41 | 1.09 | 0.10 |
|  | No (Ref) |  |  |  |  |  |
| **Asthma** | Yes | -0.25 | 0.78 | 0.41 | 1.46 | 0.43 |
|  | No (Ref) |  |  |  |  |  |
| **Thyroid** | Yes | 0.44 | 1.56 | 0.64 | 3.80 | 0.33 |
|  | No (Ref) |  |  |  |  |  |
| **Dyslipidemic** | Yes | -0.39 | 0.68 | 0.26 | 1.77 | 0.43 |
|  | No (Ref) |  |  |  |  |  |
| **CKD-ESRD** | Yes | -0.28 | 0.76 | 0.35 | 1.66 | 0.49 |
|  | No (Ref) |  |  |  |  |  |
| **IHD** | Yes | -0.17 | 0.84 | 0.44 | 1.61 | 0.60 |
|  | No (Ref) |  |  |  |  |  |
| **Neurological disease** | Yes | -2.00 | 0.14 | 0.01 | 1.24 | 0.08 |
|  | No (Ref) |  |  |  |  |  |
| **Cancer** | Yes | -1.28 | 0.2<br>8 | 0.05 | 1.51 | 0.14 |
|  | No (Ref) |  |  |  |  |  |
| **Smoking or other addiction** | Yes | 0.55 | 1.7<br>3 | 1.01 | 2.98 | 0.05 |
|  | No (Ref) |  |  |  |  |  |
| **Fever** | Yes | -0.28 | 0.7<br>6 | 0.46 | 1.24 | 0.27 |
|  | No (Ref) |  |  |  |  |  |
| **Cough** | Yes | -0.71 | 0.4<br>9 | 0.32 | 0.76 | <0.001 |
|  | No (Ref) |  |  |  |  |  |
| **Dyspnea** | Yes | -0.51 | 0.6<br>0 | 0.39 | 0.93 | 0.02 |
|  | No (Ref) |  |  |  |  |  |
| **Sore throat** | Yes | -0.79 | 0.4<br>5 | 0.29 | 0.70 | <0.001 |
|  | No (Ref) |  |  |  |  |  |
| **Asthenia** | Yes | 0.67 | 1.9<br>6 | 1.26 | 3.06 | <0.001 |
|  | No (Ref) |  |  |  |  |  |
| **Myalgia** | Yes | -0.37 | 0.6<br>9 | 0.41 | 1.16 | 0.16 |
|  | No (Ref) |  |  |  |  |  |
| **Loss appetite** | Yes | 0.33 | 1.3<br>9 | 0.83 | 2.32 | 0.21 |
|  | No (Ref) |  |  |  |  |  |
| **Loose motion** | Yes | -0.46 | 0.6<br>3 | 0.36 | 1.11 | 0.11 |
|  | No (Ref) |  |  |  |  |  |
| **Headache** | Yes | -0.45 | 0.6<br>4 | 0.33 | 1.24 | 0.19 |
|  | No (Ref) |  |  |  |  |  |
| **Nausea vomiting** | Yes | -1.24 | 0.2<br>9 | 0.13 | 0.66 | <0.001 |
|  | No (Ref) |  |  |  |  |  |
| **Nasal cold** | Yes | -0.54 | 0.5<br>8 | 0.27 | 1.24 | 0.16 |
|  | No (Ref) |  |  |  |  |  |
| **Facial pain** | Yes | -0.29 | 0.7<br>5 | 0.35 | 1.59 | 0.45 |
|  | No (Ref) |  |  |  |  |  |
| **Abdominal pain** | Yes | -0.50 | 0.6<br>1 | 0.25 | 1.48 | 0.27 |
|  | No (Ref) |  |  |  |  |  |
| **Chest pain** | Yes | -0.20 | 0.8<br>2 | 0.32 | 2.08 | 0.68 |
|  | No (Ref) |  |  |  |  |  |
| **Nasal obstruction** | Yes | -0.41 | 0.6<br>6 | 0.31 | 1.43 | 0.29 |
|  | No (Ref) |  |  |  |  |  |

## Discussion

This study evaluated the association between olfactory dysfunctions, particularly anosmia and COVID-19 infected patients’ demographic and clinical characteristics. To our knowledge, this is the first hospital-based study in Bangladesh. Out of 600 patients, about 39% patients were diagnosed with olfactory dysfunction during COVID-19 infection. Our analyses showed that patients’ age, smoking status, cough, dyspnea, sore throat, asthenia, and nausea or vomiting were significantly associated with the anosmia. We observed that fever, cough, dyspnea, sore throat, myalgia, loose motion, asthenia, and appetite loss were most common symptoms for COVID-19 infected patients.

The risk of developing anosmia for younger patients was higher than the older patients, which decreased with the increasing age. Similar finding was observed by Giacomelli et al. where they reported that OD was more frequent in younger patients.^18^ As older COVID-19 infected patients are usually more vulnerable to other comorbidities, the anosmia issue may not be recognized, and therefore this problem has not been documented frequently.^11^ However, Dong et al. showed in their study that the anosmia is more frequent in older adults. In addition, several previous studies have shown that the median age for anosmia patient was around 50 years.^19 20 21^ We found smoking or other addiction was positively correlated with Anosmia among COVID-19 patients in our study. More specifically, a smoker had double chances of having anosmia than a non-smoker during COVID-19. Al-Ani and Acharya also found similar results that smoking was significantly associated with the anosmia of the COVID-19 patients.^22^ Moreover, Katotomichelakis et al reported that smoker had six-fold risk to develop olfactory deficit than non-smokers.^23^ About (62%) patients who had comorbidity, among them almost 51% patients had Anosmia. Remarkably, Diabetic and thyroid related disorder patients had slightly more chances of Anosmia than the other patients. Though, Klopfenstein at al mention in his study, only patients with asthma had higher chances of anosmia rather than the patients with hypertension, diabetes, cardiovascular and pulmonary diseases.^24^

We noticed that, patients with asthenia correlated with the anosmia. It is clear that, the taste sensation is related with smell. Therefore, anosmia patients develop hypogeusia which may turns into ageusia too.^3 25^ Additionally, due to ageusia, the tendency of food consumption decreases among the patients and causes of weakness. A senior author Sandeep Robert Datta from Harvard Medical School mentioned that, anosmia can have serious psychological consequences during COVID-19 and it will be devastating.

Hence, weakness can be related with psychological concern like depression.^26^ Again, in the study of Lechien et. al., patients with nasal obstruction or rhinorrhea reported olfactory dysfunction, whether our study showed a negative relation between nasal obstruction and anosmia.^14 24^ Also, Nasal obstruction (49.5%) and rhinorrhea (35.0%) were frequently informed like any other complications during COVID-19 infection but not associated with olfactory dysfunctions.^24 27^ The result further indicated that, out of 232 patients with Olfactory Dysfunction, maximum patients (79) had this symptom for 4 to 8 days following to the 70 patients for almost 2 weeks which is consistent with other studies of COVID-19.^28 29^ However, 20 patients had the anosmia history for more than 2 weeks. Not surprisingly, Young patients around 3% were asymptomatic and 2% had anosmia only.

For the management of OD, two hypotheses can be applied: mucosal obstruction of the olfactory cleft causes “conductive” loss, or a post-viral anosmia syndrome with direct infection of the olfactory mucosa and destruction of the olfactory sensory neurons causes “neural” loss.^29^ These losses have different outcomes, in the conductive” loss we would expect a relatively rapid return of normal olfaction with the resolution of any other nasal symptoms. In the “neural” case, recovery would be prolonged and with a higher chance of persistent olfactory deficit.^25^ As well as the management of sudden anosmia are unknown, general guidelines can be applied for the improvement.

The total population were tested COVID-19 positive and data were taken from them directly which are true strength for our study whereas it has several limitations as well including small sample size. Due to the increased risk of contamination among staffs, we avoid direct smell identification tests and the clinical data were collected with the help of the questionnaire. As the study is hospital-based study, maximum patients were elderly and hospitalized.

## Conclusion

Our analyses show that about 39% of patients diagnosed with olfactory dysfunction. We found that patients’ age, smoking status, and asthenia are significantly positively associated with the anosmia. As anosmia can be a significant marker for the diagnosis of COVID-19, we suggest routine screening of olfactory dysfunction in patients with very early symptoms of COVID-19, particularly younger patients, smoker, and complained asthenia.

## Data Availability

We collected patients information including laboratory-confirmed COVID-19 test results from a COVID dedicated hospital, Square Hospitals Ltd., Dhaka, Bangladesh.

